# Clinical trials are moving to countries with fewer data protections

**DOI:** 10.1101/2022.07.09.22277461

**Authors:** Elad Yom-Tov, Yishai Ofran

## Abstract

**Background:** The European Union implemented data privacy laws in mid-2018 and the state of California enacted a similar law several weeks later. These regulations affect medical data collection and analysis.

**Methods:** Here we investigate the effect of these laws on clinical trials through analysis of clinical trials recorded on ClinicalTrials.gov, the World Health Organization’s International Clinical Trials Registry Platform and scientific papers describing clinical trials.

**Results:** The number of phase 1 and 2 trials in countries not adhering to data privacy laws rose significantly after implementation of these laws. The largest rise occurred in countries which are less free, as indicated by the negative correlation (−0.48, p=0.008) between the civil liberties freedom score of countries and the increase in the number of trials. This trend was not observed in countries adhering to data privacy laws nor in the paper publication record.

**Conclusions:** The implementation of data privacy laws is associated with a move of clinical trials to countries where people have fewer protections for their data.

## Introduction

The European Union’s General Data Protection Regulation (GDPR) is a regulation on data protection and privacy implemented in May 2018.^1^ In September 2018, California passed the Consumer Privacy Act (CCPA),^2^ which similarly regulates data use. The law went into effect on January 2020. These regulations attempt to provide people with stronger protections for the use of their data and to govern how medical data, among other categories, is collected, stored, and analyzed.

GDPR has special provisions which aim to enable scientific research (Article 89), but even under these provisions, significant requirements are placed on researchers.^3^ Moreover, these regulations suffer from lack of regulatory clarity and inconsistent implementation.^4,5^ Thus, in the past few years, several researchers have raised concerns about limitations on research imposed by GDPR, especially with regard to archival (secondary) use data,^6^ cross-border trials,^7^ and pediatric trials.^8^

While exploring the consequences of GDPR implementation, medical databases and personal health-related information should be divided into two different groups. One group includes data collected by electronic systems as part of the regular operation and storage protocols of health providers or insurer organizations. The other contains data collected as part of prospective clinical trials of all phases. While data of the first group are generated by routine operations and collected at the location where patients are treated, data collected from clinical trials are unique and valuable, created intentionally for a specific study. Their creation requires a laborious and expensive effort of meticulous follow-up and recording. Unlike the first group, sponsors of clinical trials are free to initiate studies and collect related data anywhere around the globe.

In a global research community, researchers from countries implementing data protection regulations could be expected to make one of two choices: either continue to conduct clinical trials in their home countries, subject to the new regulation, or move their trials to countries where such regulations do not exist or require less effort to adhere to. Thus, here we attempt to test which of these hypotheses are supported by the data. To do so, we analyzed records of clinical trials before and after the implementation of GDPR and CCPA to evaluate which of these courses the research community at large has taken. We compared those results to the scientific papers published over the same period to test if similar effects are evident in the publication record.

## Methods

We analyzed all trials registered on ClinicalTrials.gov until July 31, 2021. Even though this repository is maintained by the US government it records trials from many countries around the world and is the repository recommended by the International Committee of Medical Journal Editors^14^. As a comparison, we analyzed all trials recorded on the World Health Organization’s International Clinical Trials Registry Platform (ICTRP, https://www.who.int/clinical-trials-registry-platform, accessed June 12^th^ 2022). For the latter, the date of a trial was recorded as the enrollment date and, if that date was not provided, the registration date.

We defined the data protection implementation (DPI) date as May 25, 2018. This is the implementation date of GDPR, but as CCPA was enacted at a similar date (June 28, 2018), we chose the earlier date for all analyses. Note that since CCPA went into effect in January 2020 we expect the effect of CCPA to be delayed compared to that of GDPR.

Countries that implement GDPR include European Union members and European Economic Area members^15^. Several other countries (Andorra, Argentina, Canada, the Faroe Islands, Guernsey, Israel, the Isle of Man, Jersey, New Zealand, Switzerland, Uruguay, Japan, the United Kingdom, and South Korea)^16^ were deemed by the the EU to offer adequate levels of data protections. In the following analysis these are grouped with the GDPR-implementing countries even though they do not explicitly implement GDPR, since they may place similar limitations on data to those in GDPR-implementing countries. Separate analysis for the two groups of countries is provided in the Appendix.

We assumed that trials in the US are subject to CCPA, as many organizations have implemented CCPA for all US customers, not just those in California.^17,18^ Analysis of trials according to the US states in which they occurred is provided in the Appendix.

Trials related to COVID-19 were identified as those containing the words “COVID-19” or “SARS-COV-19” in the keywords of the ClinicalTrials data or the scientific title of the ICTRP. The phase was identified from the phase field in the ClinicalTrials and ICTRP repositories.

Additionally, we analyzed all papers which appear in the PubMed repository (https://pubmed.ncbi.nlm.nih.gov/). Papers related to COVID-19 were identified as those containing the words “COVID-19” or “SARS-COV-19” in the title or abstract. Similarly, papers identifying a trial phase were identified if a trial phase was mentioned in the title or abstract through the appearance of the phrase “phase X”, where X is 1, 2 or 3. The country affiliation of the first author and the most common affiliation of the authors were also captured.

The freedom scores of countries (specifically, civil liberties scores) were taken from Freedom House’s Global Freedom status, 2021.^19^ At the time of writing we are unaware of an agreed-upon measure for countries’ data protection. However, as protection of personal data is commonly associated with civil liberties^20^ and individual rights^21^, we used the civil liberties score from Freedom House as a proxy for data protection. See also the Appendix for additional measures provided therein.

## Results

A total of 249,387 trials were logged on ClinicalTrials.gov between January 1, 2011, and July 31, 2021. Figure 1 shows the number of new clinical trials registered per month by study phase. The number of trials in non-data protection (DP) countries (those that did not implement privacy regulations) rose more significantly after DPI (P<10^−4^) for phase 1 and 2 trials, but not for phase 3. The change was not statistically significant for GDPR countries and the US.

**Figure 1:**
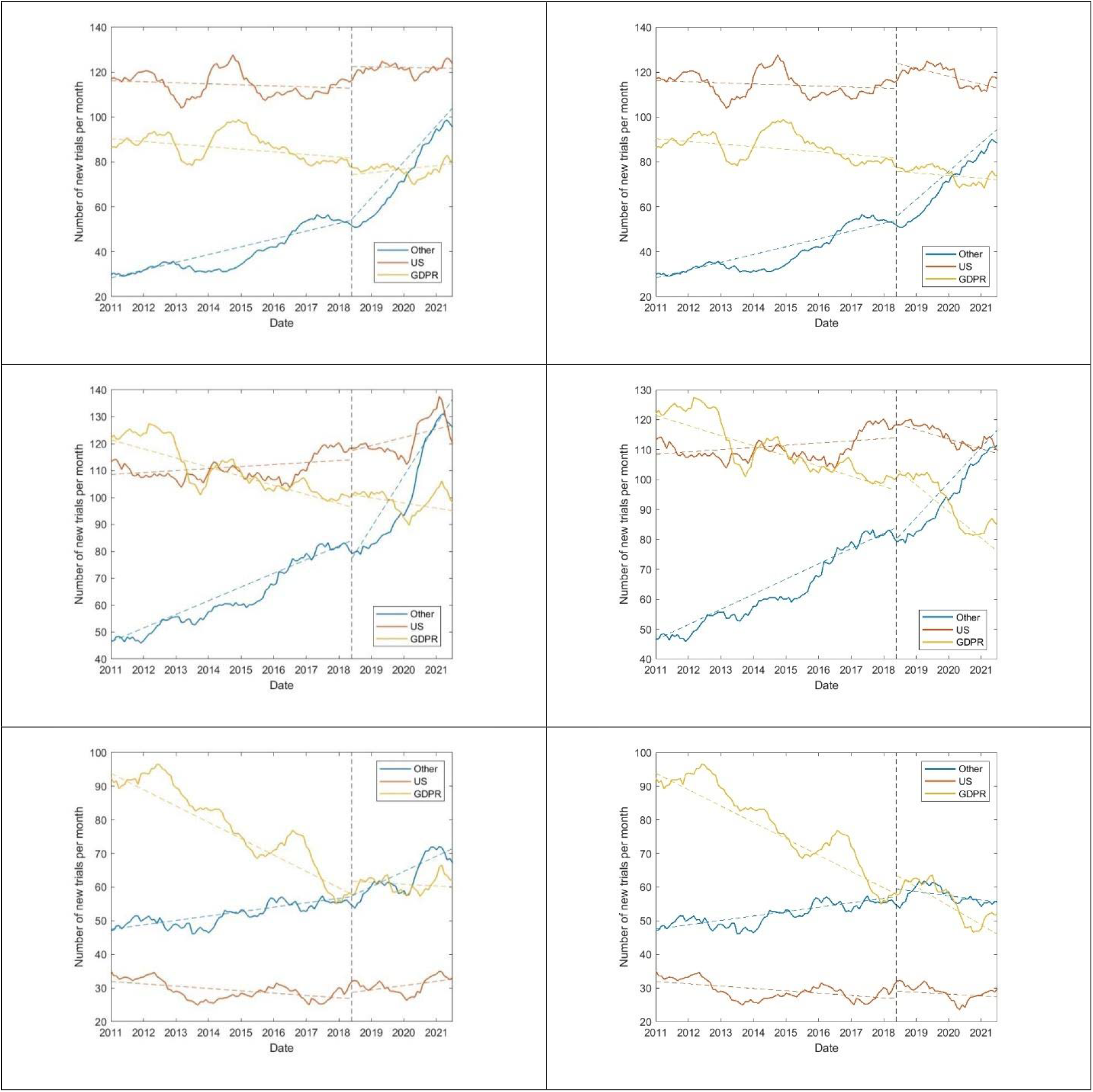
Number of new phase 1 (top), 2 (center), and 3 (bottom) clinical trials registered on ClinicalTrials.gov per month by DPI status. Graphs on the left comprise all trials, while those on the right exclude COVID-19 trials. The vertical dotted line denotes the date of DPI. Colored dotted lines are linear regression lines fit to the data.

COVID-19-related trials were most commonly phase 2 trials (49%, compared to 21% phase 1 and 30% phase 3). Among all trials the number of trials in non-DP countries rose more significantly after the data protection implementation (DPI) (P<10^−3^) for phase 1 and 2 trials, but not for phase 3. The change was not statistically significant for GDPR countries and the US.

The World Health Organization’s International Clinical Trials Registry Platform (ICTRP) logged 578,910 trials between January 1, 2011, and July 31, 2021. Figure 2 shows the number of new clinical trials registered per month by study phase. The number of trials in non-DP countries rose more significantly after DPI (P<10^−3^) for phase 1 and 2 trials. The change was not statistically significant for GDPR countries and the US for phase 1, but for phase 2 in GDPR countries and phase 3 in all countries there was a statistically significant (P<0.003) drop in the number of new trials.

**Figure 2:**
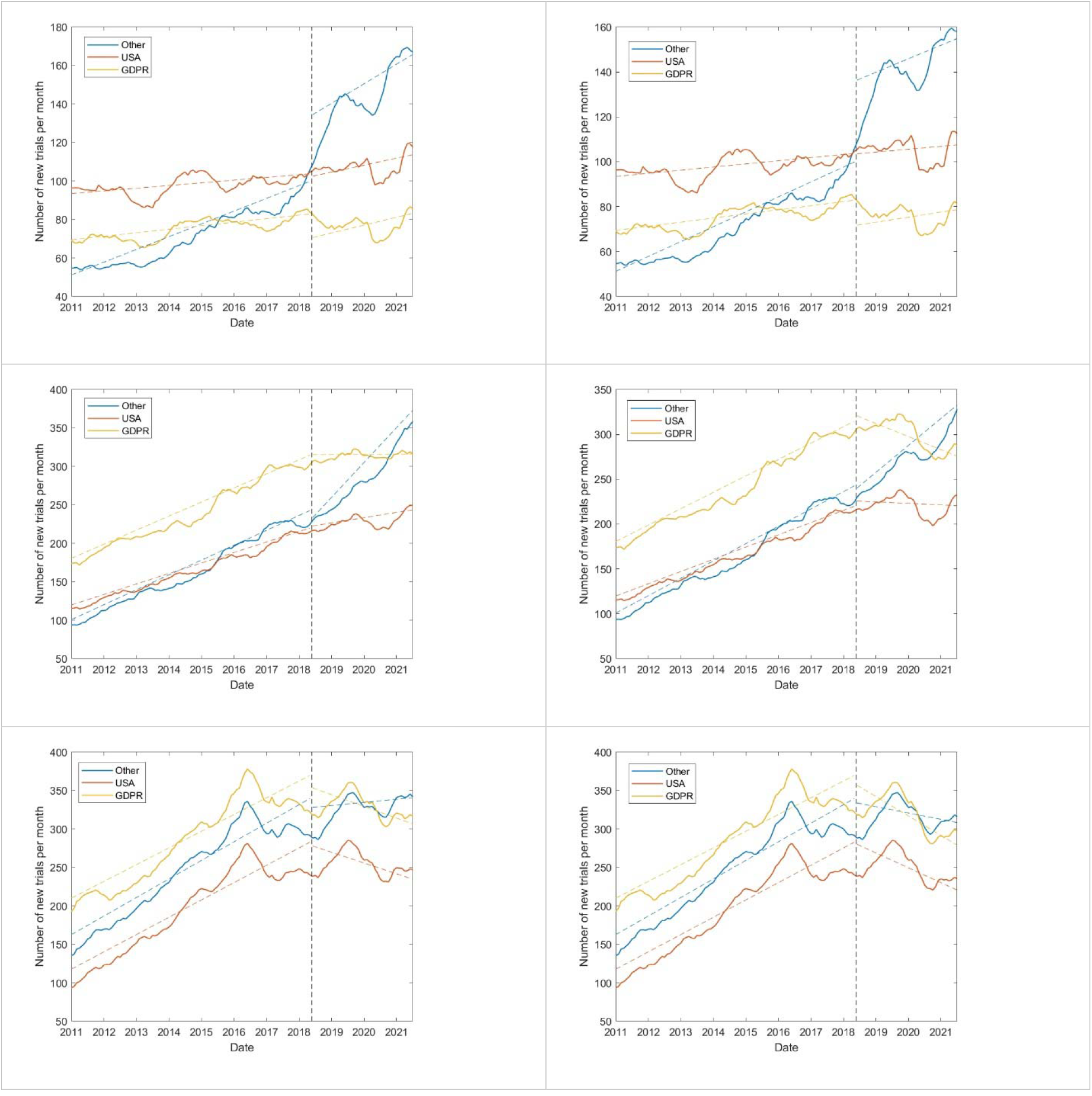
Figure 1: Number of new phase 1 (top), 2 (center), and 3 (bottom) clinical trials registered on ICTRP per month by DPI status. Graphs on the left comprise all trials, while those on the right exclude COVID-19 trials. The vertical dotted line denotes the date of DPI. Colored dotted lines are linear regression lines fit to the data.

Thus, data from ClinicalTrials.gov is consistent with that of ICTRP in that non-DP countries experienced a large rise in the number of trials after DPI, especially in phase 1 trials, whereas DP countries had no change or a drop in the number of their trials. As the changes in the number of trials over time among the two repositories is qualitatively similar, we henceforth focus on analysis of data from ClinicalTrials.gov.

Comparing the two years before DPI to the two years after, the five countries with at least 250 clinical trials that had the largest increase in the number of trials after DPI were (in descending order) Pakistan, Turkey, Mexico, Hong Kong, and Egypt (Turkey, Hong Kong, Egypt, Russia, and Taiwan at a threshold of 500 trials). The largest decrease in countries with DP occurred in Japan, South Korea, Finland, the Netherlands, and Israel (Japan, South Korea, the Netherlands, Israel, and Germany at a threshold of 500 trials). Figure 3 shows the relationship between the percentage of trials conducted in a country (out of all trials conducted in that country) after DPI as a function of the civil liberties freedom index. As the figure shows, there is a negative correlation between the freedom score of countries and the increase in the percentage of trials (−0.48, p=0.008) following DPI, indicating that the largest rise in the number of clinical trials occurred in countries which are less free. The Appendix evaluates other measures of individual freedom, the rule of law and economic freedom for their correlation with the change in the number of trials per country.

**Figure 3:**
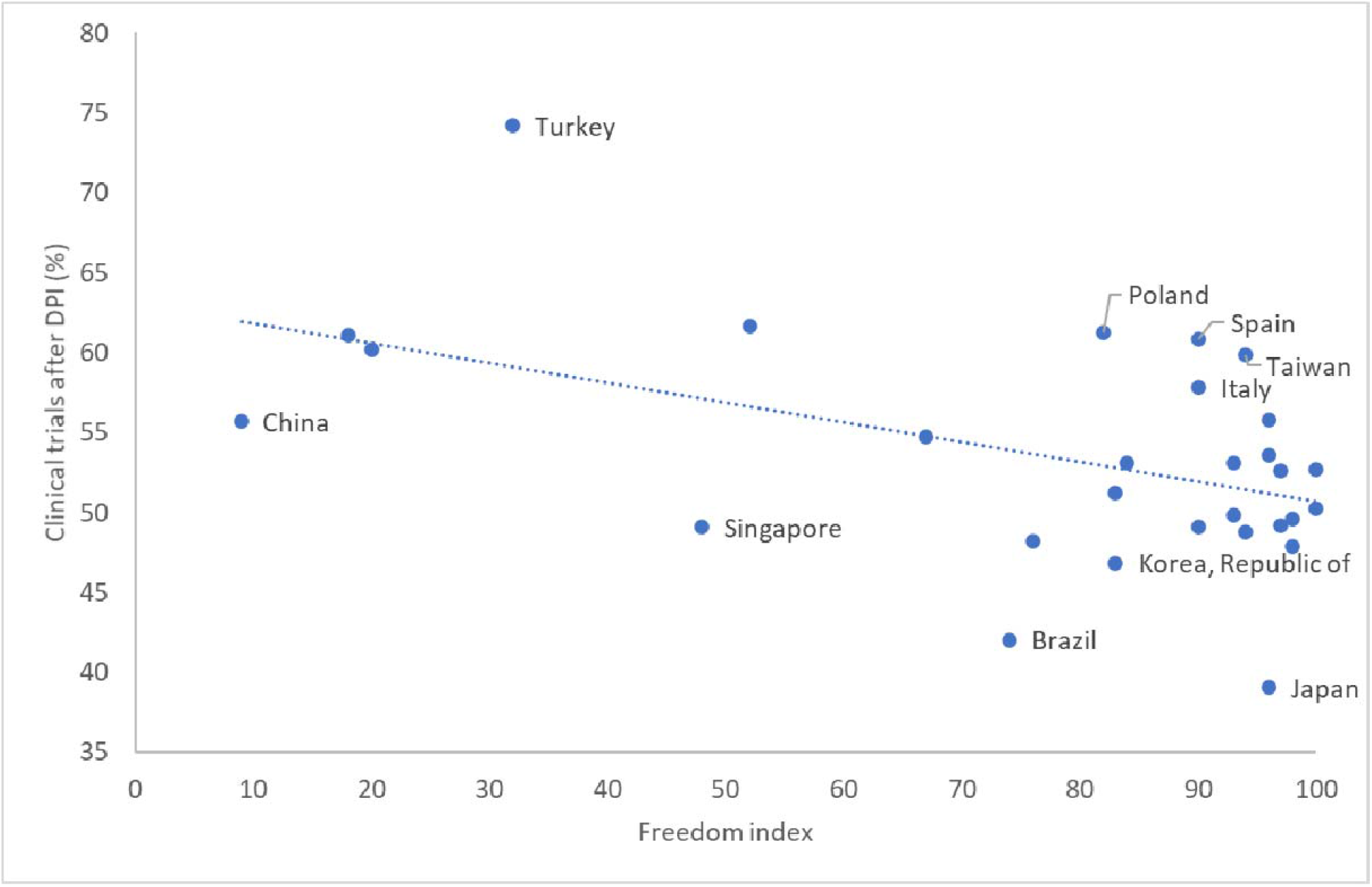
The relationship between the percentage of trials conducted after DPI (out of all trials conducted in a country) as a function of the civil liberties freedom score. The ten countries farthest from the regression line are marked.

Comparing the two years before and after DPI, the earlier the phase of a trial, the more likely it was to exhibit a larger decrease in likelihood of taking place in DP countries compared to non-DP countries following DPI (0.95, p=0.05).

Industry-funded trials increased 18% after DPI in non-DP countries but did not change in number in DP countries (chi-square test, P<10 ^-10^). Other funders (e.g., NIH, US federal) showed no similar statistically significant difference.

Both randomized and non-randomized studies increased 27% (chi-square test, P<10^−10^) in non-DP countries, while in DP countries, randomized studies increased 4% and non-randomized studies decreased by 4% (chi-square test for both, P<10^−10^).

Observational studies were less likely to occur in DP countries after DPI compared to non-DP countries (14% increase in DP countries compared to 48% increase in non-DP countries). Interventional studies had a larger relative change: 3% increase in DP countries compared to 28% increase in non-DP countries (chi-square test, P<10^−10^).

The intervention types that had the largest decrease in appearance in DP countries following DPI were behavioral, drug, and procedure. In contrast, dietary supplements, diagnostic tests, and biological products had the smallest decrease in appearance in DP countries (chi-square test, P<0.05).

Trials which were conducted in both DP and non-DP countries increased by 2% compared to a 13% rise in those which were conducted entirely within or outside DP countries (chi-square test, P=0.001).

A comparison of the average number of participants in trials after DPI compared to before it, for DP and non-DP countries, as a function of trial phase is shown in Figure 4. As the figure shows, phase 1 trials grew by approximately 13% after DPI in non-DP countries (and were reduced by 8% in DP countries), while phase 3 trials grew by 11% in DP countries.

**Figure 4:**
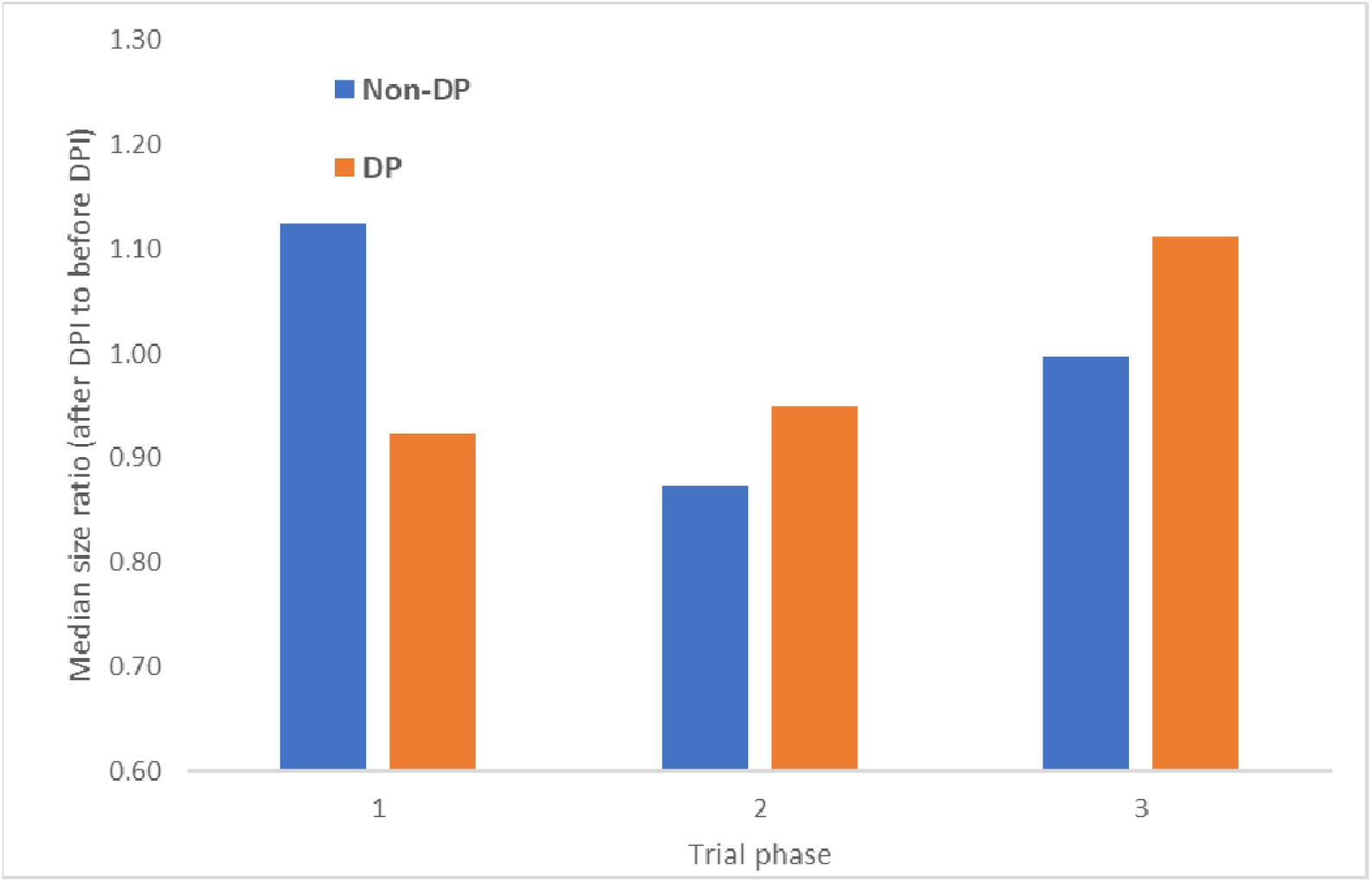
Change in the number of trial participants after DPI by trial phase and DP status.

A total of 11,078,216 papers were published on PubMed between January 1, 2011, and July 31, 2021. Of those, the trial phase was mentioned in 47,299 papers. The country affiliation of the first author and the most common affiliation were identical in 98% of papers. Therefore, we focus on the affiliation of the first author.

Figure 5 shows the number of non-COVID-19-related papers published over time, both in total and stratified by trial phase. As the figure shows, there is no discernible change in the trend of published papers following DPI.

**Figure 5:**
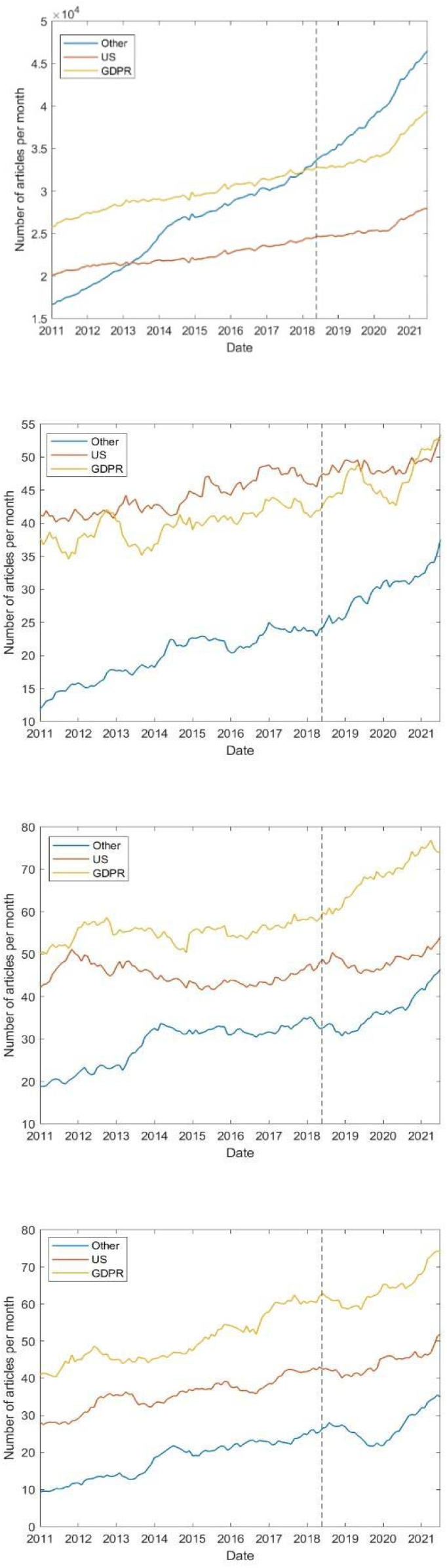
Number of papers published over time, excluding those related to COVID-19. From top to bottom: Total, phase 1, 2, and 3. The vertical dotted line denotes the date of DPI.

## Discussion

The European Union and the US state of California have attempted to regulate the use of data, thus providing more protections and choice to citizens. GDPR and CCPA have been widely adopted, potentially having a significant effect on clinical trials throughout the world. Our analysis of clinical trial registrations from two sources reveals large changes in the location, size, and type of clinical trials following the implementation of data protection laws.

The earlier the phase of a trial, the more likely it was to shift into non-DP countries following DPI. The number of phase 1 and phase 2 trials in non-DP countries rose more significantly after DPI. No similar rise occurred for phase 3 trials or in DP countries. Countries which are ranked less free experienced the largest rise in the number of clinical trials after DPI. The change in location was led by industry-funded trials, which increased significantly after DPI in non-DP countries, but did not change in DP countries. Phase 3 studies are large-scale operations that take a long time to conduct and launch; therefore, it may still be early to see the effect of DPI on phase 3 trials. Moreover, phase 3 studies require the infrastructure of multiple high-volume/high-quality centers. Long-term survival results of such studies are affected by the quality of supportive care in the community, quality of treatment for comorbidities, and logistics issues; therefore, it is a challenge to conduct phase 3 studies solely in non-DP countries. The challenges for phase 1 and 2 studies are the complexity and innovation of the technique in use. Surprisingly, we did not observe a parallel shift in the nationality of the first authors of papers following DPI; thus, the fact that industry-led studies shifted to non-DP countries probably does not reflect a scientific renaissance in these countries.

Trials relying on large amounts of data, such as observational studies, were less likely to occur in DP countries after DPI compared to non-DP countries. Similarly, intervention types that require significant data collection, including behavioral, drug, and procedure, had the largest increase in non-DP countries.Phase 1 trials in non-DP countries grew in the number of participants, whereas the size of phase 2 and 3 trials grew in DP countries.

Trials which were conducted in both DP and non-DP countries increased less than those which were conducted entirely within or outside DP countries. We attribute this to the difficulties highlighted by several researchers as to the difficulties in data sharing following the implementation of data privacy laws (see, for example, Eiss^9^, Ursin^10^, and Slokenberga^11^).

These trends in location, size, and type of trials were not reflected in the paper publication record, where there is no discernible change in the trend of published papers following DPI. This phenomenon has been described as “helicopter research”, where researchers work in low income countries with little involvement of local researchers^12^. While sponsors of clinical trials have influence on study locations decisions, there is no transparency or public access to the considerations that drive these decisions.

Taken together, our results suggest that GDPR and CCPA caused clinical trials, especially early ones and those requiring significant data collection, to be moved to countries where people have fewer protections for their data. However, the investigators remained in their original countries.

Thus, our data offer evidence for the existence of the phenomena of “ethics dumping”^12,13^, which occurs when researchers export unethical or unpalatable experiments and studies to lower-income or less-privileged settings with different ethical standards or less oversight. Interestingly, we have found that trials moved not only to lower-income countries but also to wealthy ones such as Hong Kong. This may suggest that the definition of ethics dumping should be widened to include countries where protections to individuals, whether to their privacy or to other aspects of their self, are not sufficiently rigorous.

## Data Availability

All data are publicly available from their respective sources.

## Declarations

### Author contributions

Conceptualization: EYT, YO, Methodology: EYT, YO, Analysis: EYT, Visualization: EYT, YO, Writing: EYT, YO

### Data and materials availability

All data are publicly available. The code will be deposited to GitHub after acceptance.

## References

1. European Union. Regulation (EU) 2016/679 of the European Parliament and of the council. Published online 2016. Accessed December 26, 2021. https://eur-lex.europa.eu/eli/reg/2016/679/oj

2. California Department of Justice. California Consumer Privacy Act (CCPA). Published online 2016. Accessed December 26, 2021. https://oag.ca.gov/system/files/attachments/press_releases/CCPA\%20Fact\%20Sheet\%20\%2800000002\%29.pdf

3. Staunton C, Slokenberga S, Mascalzoni D. The GDPR and the research exemption: considerations on the necessary safeguards for research biobanks. Eur J Hum Genet. 2019;27(8):1159–1167. doi:10.1038/s41431-019-0386-5

4. Mulgund P, Mulgund BP, Sharman R, Singh R. The implications of the California Consumer Privacy Act (CCPA) on healthcare organizations: Lessons learned from early compliance experiences. Health Policy and Technology. 2021;10(3):100543. doi:10.1016/j.hlpt.2021.100543

5. Becker R, Thorogood A, Ordish J, Beauvais MJS. COVID-19 Research: Navigating the European General Data Protection Regulation. J Med Internet Res. 2020;22(8):e19799. doi:10.2196/19799

6. Peloquin D, DiMaio M, Bierer B, Barnes M. Disruptive and avoidable: GDPR challenges to secondary research uses of data. Eur J Hum Genet. 2020;28(6):697–705. doi:10.1038/s41431-020-0596-x

7. Donnelly M, McDonagh M. Health Research, Consent and the GDPR Exemption. Eur J Health Law. 2019;26(2):97–119. doi:10.1163/15718093-12262427

8. Dalrymple HW. The general data protection regulation, the clinical trial regulation and some complex interplay in paediatric clinical trials. Eur J Pediatr. 2021;180(5):1371–1379. doi:10.1007/s00431-021-03933-3

9. Eiss R. Confusion over Europe’s data-protection law is stalling scientific progress. Nature. 2020;584(7822):498–498. doi:10.1038/d41586-020-02454-7

10. Ursin G. Cancer registration in the era of modern oncology and GDPR. Acta Oncologica. 2019;58(11):1547–1548. doi:10.1080/0284186X.2019.1657586

11. Slokenberga S. Biobanking between the EU and Third Countries — Can Data Sharing Be Facilitated via Soft Regulatory Tools? European Journal of Health Law. 2018;25(5):517–536. doi:10.1163/15718093-12550397

12. Nature addresses helicopter research and ethics dumping. Nature. 2022;606(7912):7–7. doi:10.1038/d41586-022-01423-6

13. Doris Schroeder, Kate Chatfield, Michelle Singh, Roger Chennells, Peter Herissone-Kelly. Equitable Research Partnerships: A Global Code of Conduct to Counter Ethics Dumping. Springer Nature; 2019.

14. De Angelis C, Drazen JM, Frizelle FA, et al. Clinical Trial Registration: A Statement from the International Committee of Medical Journal Editors. New England Journal of Medicine. 2004;351(12):1250–1251. doi:10.1056/NEJMe048225

15. Freitas M da C, Mira da Silva M. GDPR Compliance in SMEs: There is much to be done. Journal of Information Systems Engineering & Management. 2018;3(4). doi:10.20897/jisem/3941

16. Intersoft Consulting. GDPR Third Countries. Published online 2021. Accessed December 26, 2021. https://gdpr-info.eu/issues/third-countries/

17. Jedidiah Bracy. With the CCPA now in effect, will other states follow? Published online January 2, 2020. Accessed March 15, 2021. https://iapp.org/news/a/with-the-ccpa-now-in-effect-will-other-states-follow/

18. Baik JS. Data Privacy Against Innovation or Against Discrimination?: The Case of the California Consumer Privacy Act (CCPA). Social Science Research Network; 2020. Accessed June 7, 2022. https://papers.ssrn.com/abstract=3624850

19. Freedom House. Global freedom statuses. Published online 2021. Accessed December 26, 2021. https://freedomhouse.org/countries/freedom-world/scores

20. Rodotà S. Data Protection as a Fundamental Right. In: Gutwirth S, Poullet Y, De Hert p, de Terwangne c, Nouwt S, eds. Reinventing Data Protection?. Springer Netherlands; 2009:77–82. doi:10.1007/978-1-4020-9498-9_3

21. Kuner C. An international legal framework for data protection: Issues and prospects. Computer Law & Security Review. 2009;25(4):307–317. doi:10.1016/j.clsr.2009.05.001

