## Supplementary materials for "Clinical trials are moving to countries with fewer data protections"

### Comparison of ClinicalTrials.gov to the World Health Organization’s clinical trial registry

Approximately 52% of the trials in the WHO registry are also recorded in ClinicalTrials.gov, as evident by their trial number (prefix NCT). Therefore, Figure 2 shows the number of trials over time after excluding those trials, which were already analyzed in the main text.

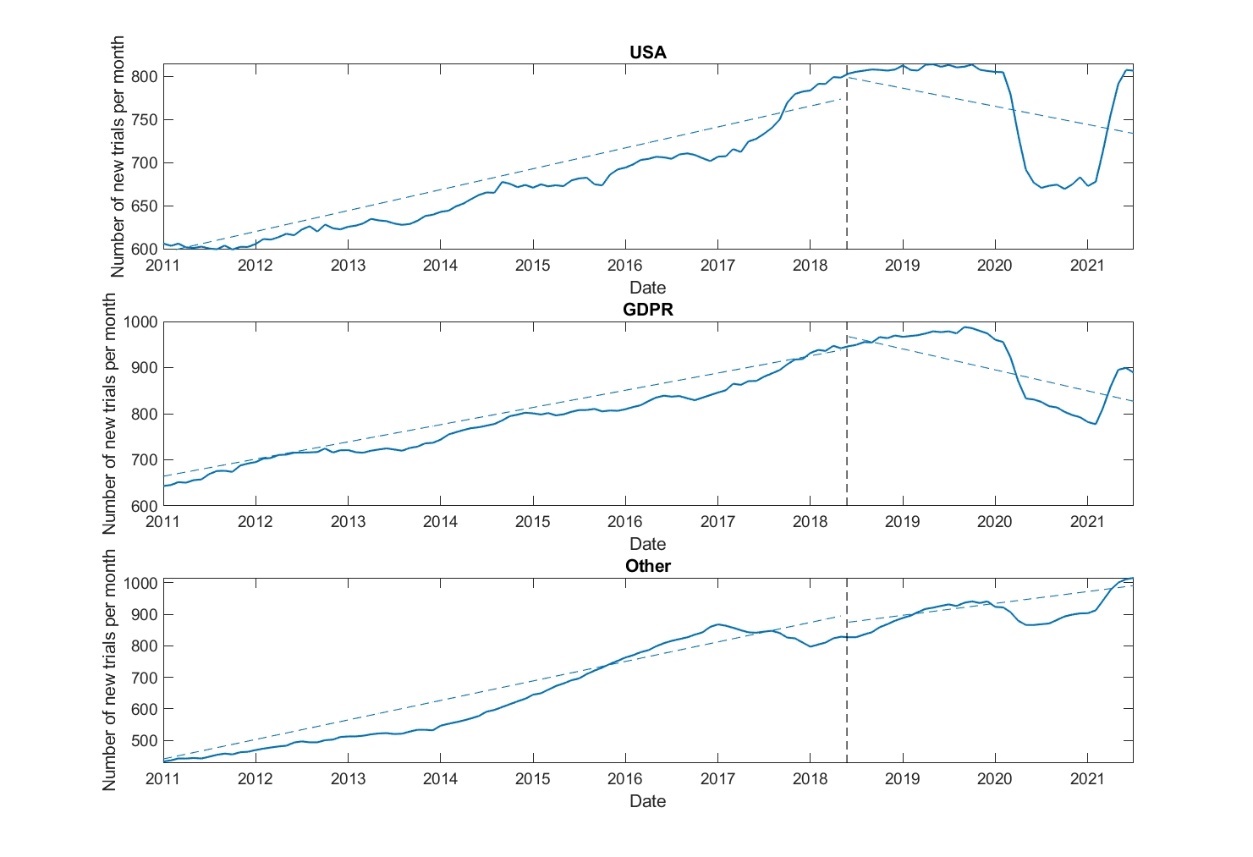

Figure 2: Number of new clinical trials in the WHO trial repository excluding trials from ClinicalTrials.gov (and excluding COVID19 trials) per month by DPI status. The vertical dotted line denotes the date of DPI. Linear regression lines fit to the data are also shown.

### EU and EEA countries versus third countries deemed to have adequate data protection under GDPR

We partitioned the countries analyzed as GDPR-implementing countries to those in the EU or EEA and those that were deemed to have adequate data protection under GDPR (hereto referred to as “other countries”). The former group includes Austria, Belgium, Bulgaria, Croatia, Cyprus, Czechia, Czech Republic, Denmark, Estonia, Finland, France, Germany, Greece, Hungary, Iceland, Ireland, Italy, Latvia, Liechtenstein, Lithuania, Luxembourg, Malta, Netherlands, Norway, Poland, Portugal, Romania, Slovakia, Slovenia, Spain, Sweden, Switzerland, and the United Kingdom. The latter group includes Andorra, Argentina, Canada, Faroe Islands, Guernsey, Israel, Isle of Man, Jersey, New Zealand, Uruguay, Japan, and South Korea.

Figure 1 compares the percentage of non-COVID19 trials registered in clinicaltrials.gov that occurred in EU and EEA countries with those in other countries. Trials in non-DP countries are shown for comparison. As the figure shows, the number of new trials per month in the EU\EEA had an increasing slope prior to GDPR implementation, after which it levelled off. There was positive slope in other countries prior to GDPR (though the slope was not as large as EU\EEA countries). After GDPR implementation the slope of new trials was negative, meaning that fewer new trials per month were registered. The change in both cases is statistically significant (P<0.05).

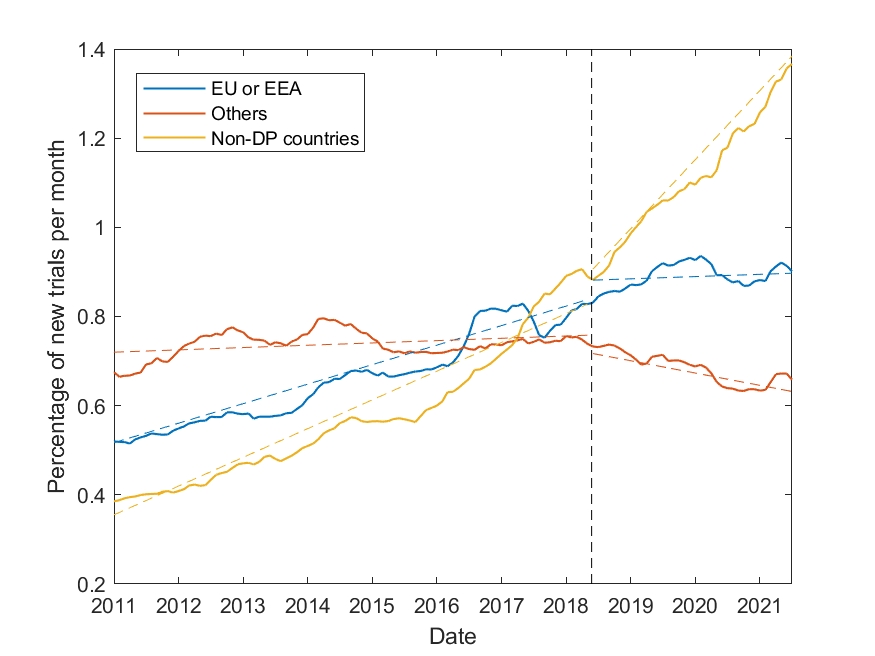

Figure 3: Percentage of new clinical trials registered in ClinicalTrials.gov in EU\EEA countries and in other countries deemed to have adequate data protection. Trials in non-DP countries are shown for comparison. Data for each line is normalized to a total of 100% to facilitate comparison.

### California versus other US states

We partitioned the data from clinicaltrials.gov about trials in the United States to three groups:

1. Trials conducted in California
2. Trials conducted in states other than California
3. Trials conducted in California and at least one other US state

Figure 2 shows the percentage of new non-COVID19 trials per month in each of the three groups of states. Trials in non-DP countries are shown for comparison. As the figure shows, prior to DPI all three groups had a positive slope, meaning that there were more new trials every month. After DPI, trials in California levelled off and those outside California had a slow negative trend after that date. Trials in both California and other states had the largest change in their trend.

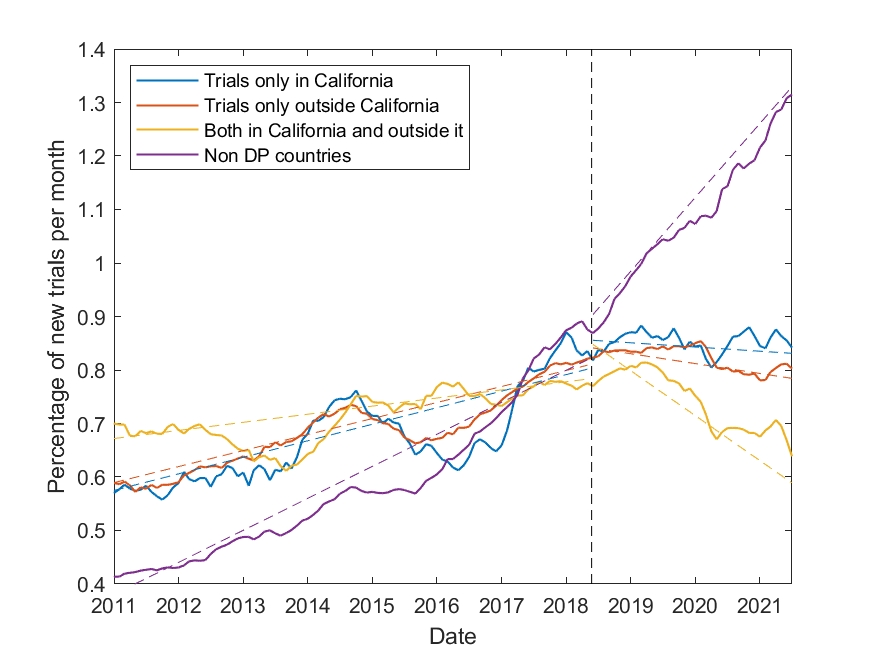

Figure 4: Percentage of new clinical trials in the US over time partitioned by state, excluding COVID19 trials. Trials in non-DP countries are shown for comparison. Data for each line is normalized to a total of 100% to facilitate comparison.

### Other measures of human freedom

We tested the correlation in the percentage of change in the number of trials per country following DPI with factors pertaining to individual freedom and economic freedom. These were taken from the Cato Institute’s 2020 Human Freedom Index (<https://www.cato.org/human-freedom-index/2020>).

The results are shown in Table 1. As the table shows, the factors most correlate with the change in the number of trials are personal freedom and the rule of law. Factors pertaining to personal relationships (marriage, etc.), trade, and property rights are less well correlated.

| Factor | Correlation | p-value |
| --- | --- | --- |
| Rule of Law | -0.47 | 0.01 |
| Personal freedom score | -0.45 | 0.02 |
| Legal System & Property Rights | -0.40 | 0.03 |
| Freedom to trade internationally | -0.23 | 0.24 |
| Identity & Relationships | -0.13 | 0.52 |

Table 1: Correlation between the change in the number of trials following DPI implementation and country-level indices (n=28).
